# Multinational modeling of SARS-CoV-2 spreading dynamics: Insights on the heterogeneity of COVID-19 transmission and its potential healthcare burden

**DOI:** 10.1101/2020.04.14.20064956

**Authors:** Erick R. Martinez-Loran, J. Jesús Naveja, Omar Y. Bello-Chavolla, Flavio F. Contreras-Torres

## Abstract

**Background:** Modelling and projections of COVID-19 using a single set of transmission parameters can be an elaborated because the application of different levels of containment measures at different stages of the worldwide COVID-19 outbreak.

**Methods:** We developed a piecewise fitting SEIR methodology to fit the progress of the COVID-19 that can be applied on any of the 185 countries listed in John Hopkins Coronavirus Resource Center. The contagious contact rate, the rate of removal and the initially exposed population were obtained at three different stages of the pandemic for a set of 18 countries, and globally for the total number of cases worldwide. The active number of infections and the removed populations were fitted simultaneously to validate the SEIR model against the available time series reports on the number of confirmed infections, recoveries and deaths. We evaluate the effect of a reduction of contagious contact rate on the level of burden put on local healthcare infrastructure considering different levels of intervention. As a guideline for future public health interventions, we also estimated the maximum number of future cases and its potential peak date.

**Findings:** We project that the peak in the number of infections worldwide will take place after the third quarter of 2020 with a decline rate that might extend beyond 2020. For 12 out of the 18 countries analyzed, we observe that, following the trend at the date of this study, the number of severe infections will surpass their healthcare capacity. For a 90% reduction scenario of the contagious contact rate, four out of the 18 countries analyzed will undergo a significant delay in the peak of infection, extending the course of the epidemic further than our simulation window (365 days).

**Interpretation:** We identify three stages for the COVID-19 transmission dynamics, which suggest that it is highly heterogeneous between countries and its contagious contact rate, is currently affected by both local responses of the public health interventions and to the population’s adherence to the measures.

**Funding:** No funding received.

## Introduction

The SARS-CoV-2 virus responsible for the coronavirus disease 2019 (COVID-19) emerged in China at the end of 2019. First reports from the World Health Organization (WHO) indicated that most of the regions with more than a thousand cases of SARS-CoV-2 infection exhibited an exponential progression in the number of cases within three weeks of the first confirmed case.^1^ A mortality rate of 3% was suggested, which has reached an alarming value of 15·2% outside of China.^2^ Notably, the mortality rate and the number of confirmed cases vary from country to country, and as of April 2020, the COVID-19 pandemic has globally reached more than 1,840,000 confirmed cases and 117,000 deaths.^3^ Initial studies have provided valuable information at the early stages of the epidemic,^4^ and preliminarily estimates showed that the different effects of governmental actions can decrease the spread to at least some extents.^5^ Social distancing and isolation of the infected population have been widely adopted measures to control the propagation of SARS-CoV-2. Such non-pharmaceutical interventions have proven to be effective in managing the epidemic in a few countries; nevertheless, limitations associated to the implementation of strict containment policies have led to difficulties in controlling the fast increase in the number of cases.^6,7^ Currently, Europe and the US report a large number of fatalities despite social distancing measures. As SARS-CoV-2 continues to spread, healthcare systems are facing a multitude of challenges at all stages of the pandemic.^8^

Analysis on the effectiveness of the interventions aimed at controlling the transmission requires adequate mathematical models to describe the SARS-CoV-2 spreading dynamics. Despite the early efforts to model the spread of COVID-19 in mainland China, the dynamics of transmission does not extrapolate seamlessly to other countries. Based on the time series of two other well-known coronavirus diseases (MERS and SARS), the estimated basic reproduction number of COVID-19, was reported between 2·24 and 3·58, which indicates that almost the totality of the population could be infected unless effective control measures are implemented.^9^ Compartmental epidemic models such as susceptible-infected-removed (SIR)^10^ and susceptible-exposed-infected-removed (SEIR) have been widely used to describe several infectious diseases.^5,9,10–14^ In particular the SEIR model has been recently used to model the progression of COVID-19 in China.^4^ While, models of a higher complexity have been recently proposed to describe the effects of social distancing and isolation,^15^ their large number of unknown parameters renders them inadequate to extract accurate information using mathematical fits to the time series of the infections. In this work, we use the susceptible-exposed-infected-removed (SEIR) compartmental model to simultaneously fit the progression of active infections and removed populations. While it is a common practice to fit only the infected compartment, the lack of consideration of the removed population might result in a fit that does not describe the actual progression of recoveries and deaths. We present a piecewise methodology that allows us to compare the progression of the epidemic at different stages of the epidemic. We identify the contagious contact rate, the removal rate and the initially exposed population at different stages of the epidemic. We use the results of the fits to project the number infections for SARS-CoV-2 within a 365-day simulation window. Using our methodology, we make comparisons on the evolution of the infectiousness for 18 countries at different stages of the epidemic. We use the extracted pandemic parameters to make relevant projections on the dynamics of the disease for the oncoming 12 months. We consider that this information can be useful to anticipate the impact of interventions to project healthcare supply needs in the short and the long terms.^16^

## Methods

### Data sources

We used the time series of the COVID-19 reported by the Johns Hopkins Coronavirus Resource Center (hereafter JHCRC), starting from January 21 to April 14, 2020.^17,18^ We selected 18 countries corresponding to ∼10% of the total number of countries currently reported in the JHCRC database. Such a particular choice was made based on the consistency of the data with the SEIR model as evaluated by *R*^*2*^, which is used as a metric of the goodness of fit. We only take into account countries with a number of confirmed cases larger than one per 100k and a number of deaths exceeding 200. Country-level data on population size was extracted from the Department of Economic and Social Affairs of the United Nations.^19^ To evaluate the burden that the pandemic lays on the country-level healthcare system, we used the number of hospital beds reported by the World Bank Database.^20^

### The SEIR model

The total population *N* is considered to be constant and is partitioned into four disjoint groups, namely: susceptible, *S*(*t*), exposed, *E*(*t*), infected, *I*(*t*), and recovered, *R*(*t*), which are all functions of time. Individuals who have had contact with an infected person are not themselves instantaneously infectious, but rather move to the exposed compartment *E*(*t*) where they develop infectious symptoms at an incubation rate *σ*.^21^ A set of coupled ordinary differential equations (ODEs) describing the spreading of the disease and the evolution of the partitions can define the SEIR model as follows:

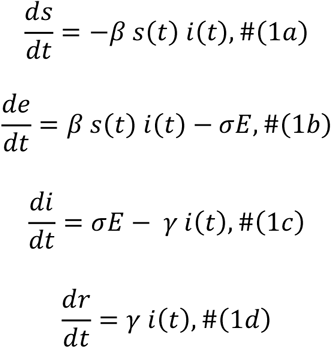

subject to the restriction

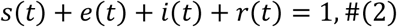

where *s*(*t*) = *S*(*t*)/*N, e*(*t*) = *E*(*t*)/*N, i*(*t*) = *I*(*t*)/*N, r*(*t*) = *R*(*t*)/*N*, the coefficients to be determined *β* ≥ 0, *γ* ≥ 0 and *σ* ≥ 0 are the effective contagious contact rate of spread, the removal (including deaths and recovered patients) rate, and the incubation rate, respectively. The time *t* is taken in days. A local epidemic scenario occurs when the number of the first infected patients increases, namely,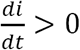. The basic reproductive number, *R* = *β*/*γ*, commonly gives the number of secondary infections generated by the first infected individual.

We solve the coupled system using SciPy’s implementation of the explicit Runge-Kutta method of order 8 (DOP853).^22^ This methodology enables us to estimate *β* and *γ*, as well as the initially exposed population *E*_0_ *≡ E*(*t* = 0) by fitting the progression of *R*(*t*) and *I*(*t*). We use the trust-reflective non-linear least-squares method to minimize the residuals between the SEIR model and the data from the time series. The values *R*(*t*) and *I*(*t*) were taken as linear combinations of the number of confirmed cases *C*(*t*), confirmed recoveries *R*_*r*_(*t*) and confirmed deaths *R*_*d*_(*t*) with *R*(*t*) = *R*_*r*_(*t*) + *R*_*d*_(*t*)and *I*(*t*) = *C*(*t*) − *R*(*t*). For purposes of following the progression at different stages of the epidemic, the values *I*_*n*_ = *I*(*t* = *t*_*n*_), *R*_*n*_ = *R*(*t* = *t*_*n*_) and *E*_*n*_ = *E*(*t* = *t*_*n*_) can be used as inputs of the SEIR model to determine the evolution of the system.

### Fitting SEIR parameters

We estimate the contagious contact rate (*β*) by fitting the data at three different stages of the COVID-19 spread. The parameter *β* is difficult to estimate accurately because the transmission rate is susceptible to changes related to government intervention. Hence, *β* can range several orders of magnitude, depending on the government response to contain the outbreak locally. Early reports from Wuhan estimated a value of 0·064 at the initial stage of the spread,^23^ but after clinical support was adopted by the Chinese government and considered in the models, it increased to 0·322. On the other hand, the removal and incubation rates (*γ* and *σ*, respectively) are not directed by population controls; according to empirical data, it was shown that these rates could range in the order of days (from 2 to 14 days).^24^ Because of this, we impose the following bounds on the possible values that these fitted parameters can take: 1/*β* ∈ [0,10], 1/*γ* ∈ [1 *day*, 1 *year*], *E*(*t* = 0) ∈ [0, *N*], and finally 1/σ= 5·1 days which is a robust estimation of the average incubation time.^25^

### Piecewise SEIR fitting and projection

We fitted *I*(*t*) and *R*(*t*) piecewise and optimized the interval ranges until the value of *R*^*2*^was higher than 0·9 (0·8 for the particular case of Mexico which is discussed in the following sections of this work). We defined the ratio of transmission rates from subsequent intervals (*Q*_*n*_ = *β*_n+1_/ *β*_*n*_), for n = 1,2,3 being the interval number. This estimate is useful to identify the changes in the COVID-19 spread dynamics upon changes in public health policies. Functional prediction bands *pb*(*t*) = *f*(*x*) ± *δ*(*x*) were obtained using *δ*(*x*) =(variance)×(inverse of the t-student cumulative distribution).

### Model projections on healthcare burden

A fundamental question for decision-makers is whether the healthcare system will be able to cope with the increasing number of patients infected with SARS-CoV-2.^26–28^ To add this practical perspective into our analysis, we considered the number of hospital beds per capita.^20^ We split the projected number of COVID-19 into three categories according to their severity: (a) mild cases that do not require hospitalization accounting for 80% of the projected infections, (b) severe disease cases which do require hospitalization, accounting for 15% of the projected infections and (c) acute disease cases, which require admission to the intensive care unit, accounting for 5% of the number of projected infections.^29^ We have performed projections for four different scenarios: one assuming the best fit of the SEIR model at the latest stage, and three scenarios where we assume a 10%, 50%, and 90% decrease in β. As a metric of the level of healthcare burden under each scenario, we define the healthcare stress as the ratio between the number of infections that require hospitalization at the peak of the disease and the number of available beds.^20^ The lethality rates were used to assess the status of the COVID-19 outbreak.

## Results

### Confirmed cases and lethality rates in different countries

The current officially reported lethality rate for COVID-19 is approximately 3·4%.^1^ Our fitted data indicates a global lethality rate of about 6·87%. Since this estimation considers only confirmed cases, we argue that the former value could be an overestimation introduced by underreporting the actual number of infections. A cluster of European countries (UK, Italy, Spain and France) whose calculated lethality rate range from 10·3 to 12·7% and their number of confirmed cases exceeds 100 cases per 100k encompasses the most affected countries (figure 1). Germany, Austria, Switzerland and the US reported >100 cases per 100k and their lethality rate is similar or less than that of China (4·02%). South Korea, Singapore, Australia and Canada also can be clustered together, being countries with a lower lethality rate (< 3%) and confirmed cases less than 100 per 100k. Brazil, Ecuador and Iran have lethality rates higher than China and their number of confirmed cases is currently increasing (>10 per 100k). Finally, Japan and Mexico show a number of confirmed cases less than 10 per 100k. The lethality rate for Mexico (6·47%) is higher than that of China, however, in this study, Mexico shows the lowest number of confirmed cases per 100k habitants. Such a comparison is only a geographical indication that the epidemic has different dynamics of transmissibility.

**Figure 1.**
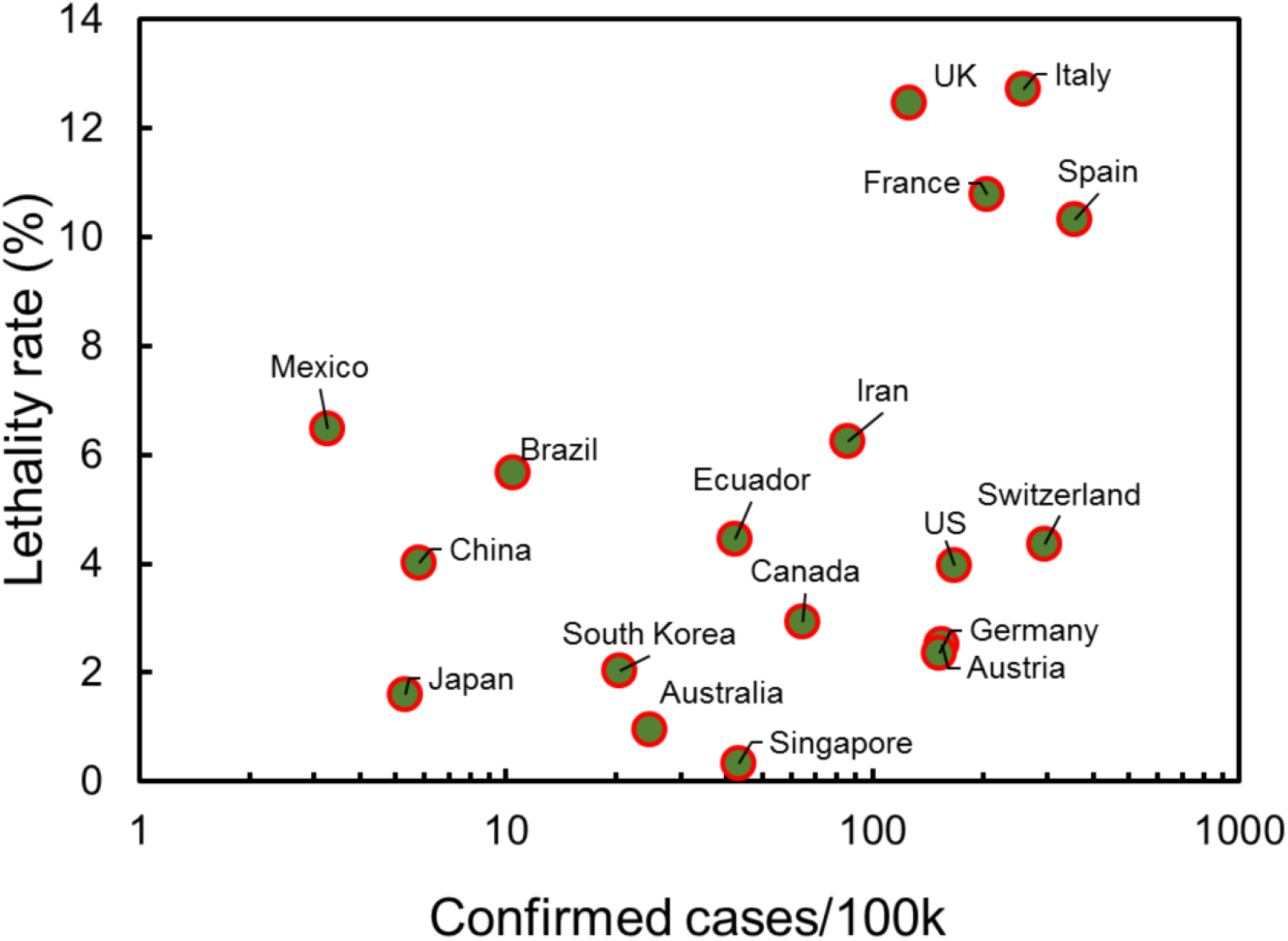
Number of confirmed COVID-19 cases per 100k habitants versus the lethality rate (in %) for different countries. The lethality rate was calculated as the quotient of deaths by COVID-19 divided by the confirmed cases according to the data published from JHCRC, on April 14, 2020.

### Differences in SEIR model fitting across countries

In figure 2b, the time series of the active infections and removed individuals shows an effective change in both contagious contact and removal rates, leading to a crossover in February 15 with a transitory decline in the number of active infections. A second cross-point around March 21 is observed, when the number of active infections takes over the number of recoveries. These features can be interpreted to be due to a lag between the pandemic’s onsets across different regions of the world. Such timing differences introduce an additional heterogeneity due to local reporting and containment policies. To address the heterogeneity of the COVID-19 outbreak, the SEIR model is used to model the progression of COVID-19 in 18 countries (see Supplementary Material). The crossing point between the infected and removed curves is currently missing on all of the countries studied, with a clear exception for China, South Korea and Austria. Because of the inherently high effectiveness of the transmission of SARS-CoV-2, it is expected that the epidemic will extend for several months in some countries. According to the current trend, we project that the peak in the number of infections worldwide will take place after the third quarter of 2020 (figure 2c). We estimate that a 90% reduction in the contagious contact rate (0.1*β*) will be required to bring the peak of on the number of infections to a date within the second quarter of 2020 (figure 2d).

**Figure 2.**
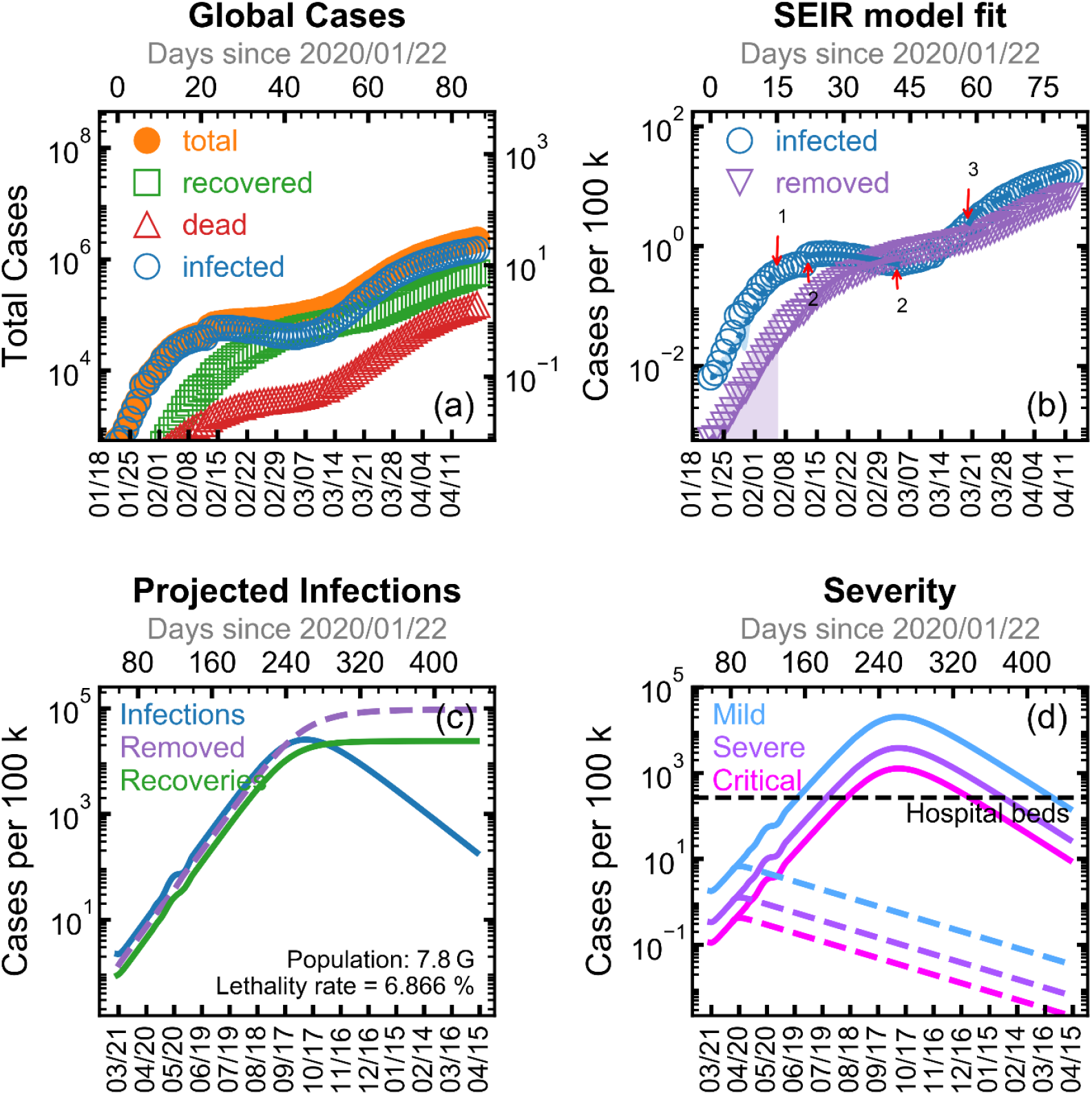
Fit of the SEIR model to worldwide data. In all cases, the solid lines represent to our data fitted, while markers correspond to official data. The *x*-axis is the period of time in format mm/dd. Upper left: total infections, recoveries, deaths and infected cases as per official reports from JHCRC. Figure 2a shows the global picture of the progress of COVID-19 epidemic as reported by JHCRC, including the SEIR model fit and confidence intervals for both infected and removed (shadow intervals indicate the piecewise fitting for obtaining the effective infectious contact rate of spread for each period). In solid orange circles is the total number of confirmed cases per day *C*(*t*), the number of confirmed recoveries *R*_*r*_(*t*) and deaths *R*_*d*_(*t*) are indicated in open green squares and open red triangles respectively. The active number of infections at time *t* estimated as *I*(*t*) = *C*(*t*) − *R*_*r*_(*t*) − *R*_*d*_(*r*) is indicated by open blue circles. Figure 2b shows the selection of intervals used to fit the mode globally: (a) from the date of the first detected case to the point 1 indicated on the infections curve, (b) on the range indicated between the arrows labeled as 2 on the infections curve and (c) from the point labeled as 3 on the infections curve to April 14, 2020. Figure 2c indicates the projections accordingly the best-fit model for infections, removed and recovery cases. Lethality rate is shown for comparison. Figure 2d indicates the severity of the spread, in which solid lines represent splitting the projected curve into severity categories, whereas dashed lines represent the projection with 90% reduction of current contagious contact rate.

So far, European countries still lead in the number of confirmed cases and COVID-19 deaths per capita, followed by the United States (see Table 2). With the exception of Japan and Mexico, it seems that other countries studied have managed to slow down the spreading of the epidemic as revealed by lower *β*_3_ and *Q*_1,2_ < 1 values (figure 3). In the case of Mexico, the goodness of fit is limited (*R*^*2*^ > 0.8) due to a different sampling methodology. Namely, as a response to AH1N1 2009 outbreak in Mexico, the healthcare system implemented a proper sentinel surveillance model, which samples only a few of all SARS-CoV-2 infections;^30^ therefore, the effect of public policy measures for this country might not be accurately estimated using our predictions. Substantial declines in spreading rates were observed for Austria, China and South Korea. Notably, Austria was an early responder of the epidemic spread by enforcing more stringent restrictions, and thus it seems that is preparing to lift lockdown.^31^ Removal rates (associated to both: deaths and recoveries in figure 2c) appear to be increasing in all countries, which goes along with a rise in the estimations of lethality rates, and it might also be related with an effective increase in the recovery rates as available data on the progress of the epidemic becomes available.

**Table 1.** Piecewise analysis for each evaluated country using the SEIR model. The parameters model to be determined, namely, *β*≥0, *γ*≥0 and *e*≥0 are the effective contagious contact rate of spread, the removal (including deaths and recovered patients) rate, and the initially exposed population, respectively. The fitted values stand for three different stages for the COVID-19 spread. The goodness of fit, R^2^, is used to demonstrate the consistency of SEIR model with the time series (from the Johns Hopkins Coronavirus Resource Center) as from January 21 to April 14, 2020. The ratio Q’s of the transmission rate (no units) is calculate as *Q*_*n*_=β_n+1_ / β_*n*_ (with n=1, 2, and 3 stages). *Q*_*n*_ < 1 implies the effectiveness in the containment measures for each country (i.e. local cases) over the different stages of the epidemic. All countries are listed in alphabetic order. The Global case represent the fit for all the 185 countries listed in the database from Johns Hopkins Coronavirus Resource Center, by April 14, 2020.

|  | First stage |  |  |  | Second stage |  |  |  | Third stage |  |  |  |  |  |
| --- | --- | --- | --- | --- | --- | --- | --- | --- | --- | --- | --- | --- | --- | --- |
| Country | $\beta_1$ | $\gamma_1$ | $e_1$ | $R^2$ | $\beta_2$ | $\gamma_2$ | $e_2$ | $R^2$ | $\beta_3$ | $\gamma_3$ | $e_3$ | $R^2$ | $Q_1$ | $Q_2$ |
| Australia | 0.055<br>(0.039-0.077) | 0.018<br>(0.014-0.023) | 7<br>(6-8) | 0.973 | 0.420<br>(0.419-0.420) | 0.011<br>(0.011-0.011) | 25<br>(25-25) | 0.995 | 0.006<br>(0.001-0.023) | 0.021<br>(0.019-0.023) | 3229<br>(2978-3501) | 0.993 | 7.67 | 0.01 |
| Austria | 0.746<br>(0.719-0.774) | 0.005<br>(0.002-0.009) | 7<br>(6-8) | 0.999 | 0.329<br>(0.328-0.329) | 0.003<br>(0.003-0.003) | 1156<br>(1154-1158) | 0.996 | 0.000<br>(0.000-inf) | 0.043<br>(0.040-0.046) | 6534<br>(5863-7281) | 0.974 | 0.44 | 3.9E-10 |
| Brazil | 0.660<br>(0.586-0.744) | 0.003<br>(0.000-0.095) | 2<br>(1-3) | 0.990 | 0.912<br>(0.905-0.919) | 0.006<br>(0.005-0.007) | 55<br>(50-59) | 0.996 | 0.183<br>(0.172-0.194) | 0.008<br>(0.006-0.011) | 2452<br>(2143-2806) | 0.995 | 1.38 | 0.20 |
| Canada | 0.031<br>(0.023-0.042) | 0.014<br>(0.011-0.017) | 5<br>(4-5) | 0.937 | 0.521<br>(0.518-0.524) | 0.005<br>(0.004-0.005) | 0<br>(0-3.15E7) | 0.986 | 0.143<br>(0.129-0.159) | 0.039<br>(0.036-0.043) | 5875<br>(5213-6622) | 0.986 | 16.75 | 0.27 |
| China | 0.469<br>(0.425-0.518) | 0.013<br>(0.008-0.022) | 3441<br>(2902-4079) | 0.993 | 0.000<br>(0.000-inf) | 0.033<br>(0.033-0.034) | 38804<br>(38196-39421) | 0.984 | 0.000<br>(0.000-0.000) | 0.082<br>(0.082-0.083) | 3995<br>(3785-4215) | 0.999 | 2.1E-14 | 7.5E-14 |
| Ecuador | 0.042<br>(0.023-0.079) | 0.003<br>(0.000-0.017) | 9<br>(8-11) | 0.990 | 0.451<br>(0.449-0.453) | 0.006<br>(0.006-0.006) | 526<br>(524-528) | 0.972 | 0.161<br>(0.138-0.187) | 0.013<br>(0.009-0.019) | 758<br>(492-1168) | 0.979 | 10.67 | 0.35 |
| France | 0.026<br>(0.015-0.046) | 0.022<br>(0.018-0.027) | 6<br>(5-8) | 0.824 | 0.579<br>(0.578-0.580) | 0.005<br>(0.005-0.005) | 171<br>(171-172) | 0.991 | 0.144<br>(0.130-0.159) | 0.047<br>(0.043-0.051) | 15519<br>(13280-18135) | 0.965 | 22.06 | 0.24 |
| Germany | 0.029<br>(0.018-0.046) | 0.003<br>(0.001-0.010) | 12<br>(11-13) | 0.983 | 0.446<br>(0.446-0.446) | 0.003<br>(0.003-0.003) | 400<br>(400-400) | 0.991 | 0.075<br>(0.063-0.091) | 0.050<br>(0.047-0.054) | 43680<br>(38319-49790) | 0.963 | 15.51 | 0.16 |
| Iran | 2.084<br>(1.867-2.325) | 0.115<br>(0.099-0.133) | 27<br>(18-42) | 0.994 | 0.166<br>(0.165-0.166) | 0.055<br>(0.055-0.055) | 4920<br>(4908-4933) | 0.987 | 0.012<br>(0.000-2.456) | 0.115<br>(0.108-0.124) | 15315<br>(9980-23502) | 0.976 | 0.07 | 0.07 |
| Italy | 0.011<br>(0.004-0.032) | 0.003<br>(0.001-0.008) | 1<br>(1-1) | 0.982 | 0.415<br>(0.415-0.415) | 0.035<br>(0.035-0.035) | 947<br>(947-947) | 0.995 | 0.064<br>(0.060-0.068) | 0.027<br>(0.026-0.028) | 42634<br>(40231-45182) | 0.992 | 37.20 | 0.15 |
| Japan | 0.166<br>(0.121-0.227) | 0.007<br>(0.002-0.031) | 7<br>(5-10) | 0.936 | 0.140<br>(0.139-0.141) | 0.019<br>(0.019-0.019) | 92<br>(91-92) | 0.994 | 0.180<br>(0.176-0.184) | 0.016<br>(0.014-0.018) | 0<br>(0-inf) | 0.994 | 0.84 | 1.28 |
| Mexico | 0.000<br>(0.000-0.000) | 0.034<br>(0.019-0.062) | 7<br>(6-8) | 0.883 | 0.280<br>(0.279-0.280) | 0.007<br>(0.007-0.007) | 134<br>(134-135) | 0.993 | 0.321<br>(0.180-0.571) | 0.084<br>(0.061-0.115) | 245<br>(4-15573) | 0.801 | 1.7E+15 | 1.14 |
| Singapore | 0.122<br>(0.101-0.147) | 0.036<br>(0.030-0.043) | 19<br>(16-23) | 0.912 | 0.193<br>(0.031-1.207) | 0.003<br>(0.000-15734.2) | 31<br>(6-149) | 0.975 | 0.158<br>(0.151-0.166) | 0.024<br>(0.023-0.026) | 103<br>(88-122) | 0.992 | 1.58 | 0.81 |
| South Korea | 0.100<br>(0.072-0.138) | 0.015<br>(0.009-0.026) | 10<br>(8-13) | 0.898 | 0.238<br>(0.238-0.238) | 0.003<br>(0.003-0.003) | 1888<br>(1885-1892) | 0.979 | 0.004<br>(0.001-0.015) | 0.044<br>(0.042-0.045) | 1669<br>(1244-2240) | 0.963 | 2.38 | 0.01 |
| Spain | 0.119<br>(0.095-0.150) | 0.025<br>(0.017-0.037) | 0<br>(0-1) | 0.920 | 0.524<br>(0.524-0.524) | 0.029<br>(0.029-0.029) | 399<br>(399-399) | 0.988 | 0.072<br>(0.067-0.077) | 0.055<br>(0.054-0.056) | 49072<br>(46722-51540) | 0.996 | 4.39 | 0.13 |
| Switzerland | 0.633<br>(0.545-0.736) | 0.004<br>(0.000-0.081) | 43<br>(34-56) | 0.985 | 0.349<br>(0.346-0.352) | 0.005<br>(0.004-0.005) | 1597<br>(1575-1620) | 0.995 | 0.031<br>(0.020-0.047) | 0.050<br>(0.047-0.054) | 9546<br>(8239-11060) | 0.966 | 0.55 | 0.08 |
| United Kingdom | 0.253<br>(0.173-0.370) | 0.013<br>(0.004-0.045) | 0<br>(0-inf) | 0.910 | 0.546<br>(0.546-0.547) | 0.014<br>(0.014-0.014) | 35<br>(35-35) | 0.994 | 0.167<br>(0.157-0.177) | 0.016<br>(0.014-0.018) | 12290<br>(11112-13595) | 0.995 | 2.15 | 0.30 |
| United States | 0.026<br>(0.019-0.037) | 0.016<br>(0.013-0.019) | 8<br>(7-9) | 0.924 | 0.686<br>(0.686-0.686) | 0.005<br>(0.005-0.005) | 73<br>(73-73) | 0.996 | 0.133<br>(0.124-0.143) | 0.012<br>(0.010-0.014) | 134668<br>(124444-145731) | 0.996 | 26.05 | 0.19 |
| Global | 0.866<br>(0.768-0.976) | 0.017<br>(0.002-0.121) | 0<br>(0-inf) | 0.907 | 0.003<br>(0.000-0.024) | 0.040<br>(0.038-0.041) | 25092<br>(21822-28852) | 0.979 | 0.122<br>(0.115-0.129) | 0.028<br>(0.026-0.030) | 330024<br>(304106-358151) | 0.993 | 0.0039 | 35.79 |

**Table 2.** Piecewise analysis for each evaluated country using the SEIR model. Recovery and lethality rates (in %) are shown by April 14, 2020. The effective contagious contact rate of spread (β) was calculated for the latest stage to predict the date to which the maximum of infectious for each country. Percentages of reduction in the current transmission rate was defined at the 10% (or 0.9*β_latest_), 50% (or 0.5*β_latest_), and 90% (or 0.1*β_latest_) to measure the saturation of the available healthcare infrastructure. We observe that a 50% reduction could compromise a saturation of the healthcare capacity for several countries. The latter can suggest stricter measures of governmental actions in those countries. These projections can be related to the number of beds available for attending infectious cases. The number of hospital beds and acute care beds based on data from the World Bank Database. The peak date of maximum infectious cases is projected for each scenario. The Global case represent the fit for all the 185 countries listed in the database from Johns Hopkins Coronavirus Resource Center, by April 14, 2020.

| | Recovery (%) | Lethality (%) | Peak date $\beta$ | Peak Cases (/ 100k) | Peak date 0.9 $\beta$ | Peak Cases 0.9 $\beta$ (/100k) | Peak date 0.5 $\beta$ | Peak Cases 0.5 $\beta$ (/ 100k) | Peak date 0.1 $\beta$ | Peak Cases 0.1 $\beta$ (/ 100k) | Beds (/100 k) |
| --- | --- | --- | --- | --- | --- | --- | --- | --- | --- | --- | --- |
| Australia | 58.39 | 1.01 | 26/03/2020 | 10 | 23/04/2020 | 17 | 22/04/2020 | 17 | 22/04/2020 | 17 | 380 |
| Austria | 66.49 | 2.95 | 07/04/2020 | 84 | 17/04/2020 | 108 | 17/04/2020 | 108 | 17/04/2020 | 108 | 760 |
| Brazil | 41.64 | 6.36 | 04/08/2020 | 81992 | 13/08/2020 | 75884 | 23/10/2020 | 66214 | 13/04/2021 | 176 | 220 |
| Canada | 32.14 | 4.13 | 07/08/2020 | 82339 | 04/09/2020 | 27544 | 04/02/2021 | 10136 | 17/04/2020 | 34 | 270 |
| China | 92.59 | 5.53 | 03/03/2020 | 4 | 12/04/2020 | 1 | 12/04/2020 | 1 | 12/04/2020 | 1 | 420 |
| Ecuador | 9.92 | 4.98 | 25/07/2020 | 80154 | 04/08/2020 | 65029 | 14/10/2020 | 51665 | 12/04/2021 | 124 | 150 |
| France | 23.47 | 12.54 | 26/07/2020 | 81086 | 26/08/2020 | 21711 | 15/02/2021 | 5688 | 13/04/2020 | 76 | 650 |
| Germany | 58.78 | 3.08 | 07/10/2020 | 82944 | 10/04/2021 | 2978 | 20/04/2020 | 96 | 16/04/2020 | 90 | 830 |
| Iran | 68.01 | 6.24 | 05/04/2020 | 42 | 12/04/2020 | 33 | 12/04/2020 | 33 | 12/04/2020 | 33 | 150 |
| Italy | 24.78 | 13.19 | 31/01/2021 | 10825 | 10/01/2021 | 15432 | 13/04/2021 | 482 | 22/04/2020 | 103 | 340 |
| Japan | 9.55 | 1.94 | 07/08/2020 | 58826 | 18/08/2020 | 61802 | 08/11/2020 | 48096 | 12/04/2021 | 9 | 1340 |
| Mexico | 33.75 | 7.72 | 30/06/2020 | 83944 | 24/07/2020 | 23915 | 10/11/2020 | 9540 | 12/04/2020 | 4 | 150 |
| Singapore | 14.02 | 0.22 | 31/07/2020 | 89372 | 11/08/2020 | 46129 | 09/11/2020 | 29079 | 12/04/2020 | 37 | 240 |
| Republic of Korea | 73.62 | 2.16 | 12/04/2021 | 35 | 12/04/2020 | 19 | 12/04/2020 | 19 | 12/04/2020 | 19 | 1150 |
| Spain | 39.19 | 10.48 | 01/11/2020 | 19486 | 12/04/2021 | 1016 | 18/04/2020 | 178 | 16/04/2020 | 169 | 300 |
| Switzerland | 60.57 | 4.90 | 26/03/2021 | 53892 | 18/04/2020 | 205 | 16/04/2020 | 201 | 16/04/2020 | 197 | 470 |
| United Kingdom | 0.36 | 13.31 | 20/07/2020 | 64524 | 29/07/2020 | 59793 | 05/10/2020 | 45371 | 12/04/2021 | 72 | 280 |
| United States | 8.37 | 5.26 | 04/08/2020 | 80425 | 11/08/2020 | 63224 | 25/10/2020 | 48897 | 12/04/2021 | 169 | 290 |
| Global | 25.37 | 6.87 | 08/10/2020 | 25429 | 09/10/2020 | 34247 | 01/04/2021 | 15897 | 20/04/2020 | 8 | 270 |

**Figure 3.**
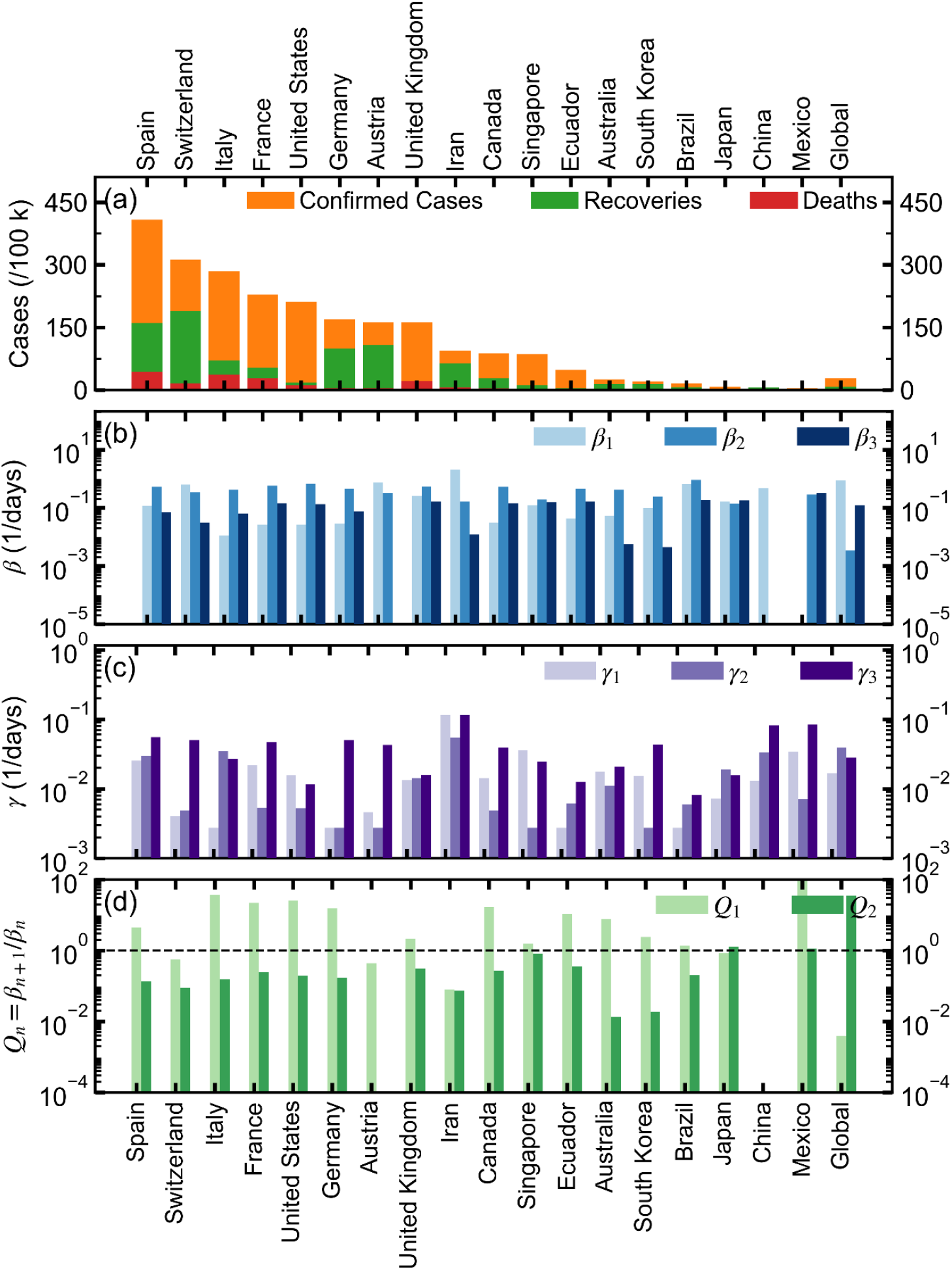
Comparison of the fitted values for the SEIR model for 18 countries. Figure 3a shows the total confirmed cases, recoveries and deaths per 100k, colored in orange, green and red, respectively. Figure 3b shows the fitted contagious contact rate (*β*) for each of the different intervals. Figure 3c shows the fitted removal rate (*γ*) for each of the different intervals. Figure 3d shows the ratio *Q*_*n*_ = *β*_*n*+1_/*β*_*n*_ between the different intervals, which is proposed as a measure of the effectiveness of the containment measures. For panels (b-d) the missing values indicate that only two intervals were identified in the corresponding dataset. The scale of the vertical axis in (d) was adjusted to a maximum of only 100. Nonetheless, Mexico overpasses this limit drastically in *Q*_2_, which could be attributed to a low early detection rate due to the implementation of a sentinel model for epidemiological surveillance (see Discussion and Table 1).

Two distinct values of *Q* are defined per country to reflect changes in their contagious contact rate for the three fitted intervals (figure 3d). Values of *Q* above the dashed line indicate an increase of *β* from interval n to interval n + 1, whereas values below that threshold indicate a reduction in the transmission rate. *Q*_1_ gives a metric of how susceptible the contagious contact rate is to updates in the reported number of cases, as more information becomes available since the early stages of COVID-19 spread for each country. Except for Switzerland, Austria, Iran China and Japan, all other studied countries show *Q*_1_ > 1. Overall, the projections show that *Q*_2_ < 1 with the exception for Japan that recently reported a surge in new cases; and Mexico, which has recently entered the exponential phase of the epidemic.

### Projections on the healthcare burden and effects of containment measures

Assuming that treatment and prevention of the disease do not have an immediate impact on the parameters of the model, our projections indicate that tighter measures should be taken to effectively reduce the spread of COVID-19. Our projections also suggest that stringent measures are required in most countries; with the only exceptions of Australia, Austria, China, Iran and South Korea, which could either continue with their ongoing policies or, up to some safe extent, relax their local interventions (figure 4). As expected, a monotonic decrease in the number of cases at the peak of the infection *I*_*peak*_ is observed with a decrease in the value of *β* (see figure 4b). For 12 out of the 18 countries analyzed (figure 4c), the current trend indicates that the number of severe infections will surpass the healthcare capacity (see details in Table 2). According to our projections, Germany, South Korea, Spain and Switzerland would avoid a saturation of the healthcare capacity if they will apply only with a 10% reduction in their current transmission rate (0.9*β*_*latest*_). We observe that for four out of the 18 countries analyzed, a 50% reduction (0.5*β*_*latest*_) is enough to avoid the saturation of the available health care infrastructure. We also see that a reduction of 90% (0.1*β*_*latest*_) will be a failsafe intervention for all the analyzed countries. For such a level of intervention, it is expected that 11 out of 18 countries (not including Australia, Austria, China and South Korea, which seem to have already passed their infection peaks) will reach their peak before the third quarter of 2020. Additionally, it is observed that five out of the 18 analyzed countries will significantly delay the peak of the infection even at 90% reduction. For such cases, the course of the epidemic will be extended further beyond the 365-day simulation window. The countries that follow the former trend are Brazil, Ecuador, Japan, the United Kingdom and the United States.

**Figure 4.**
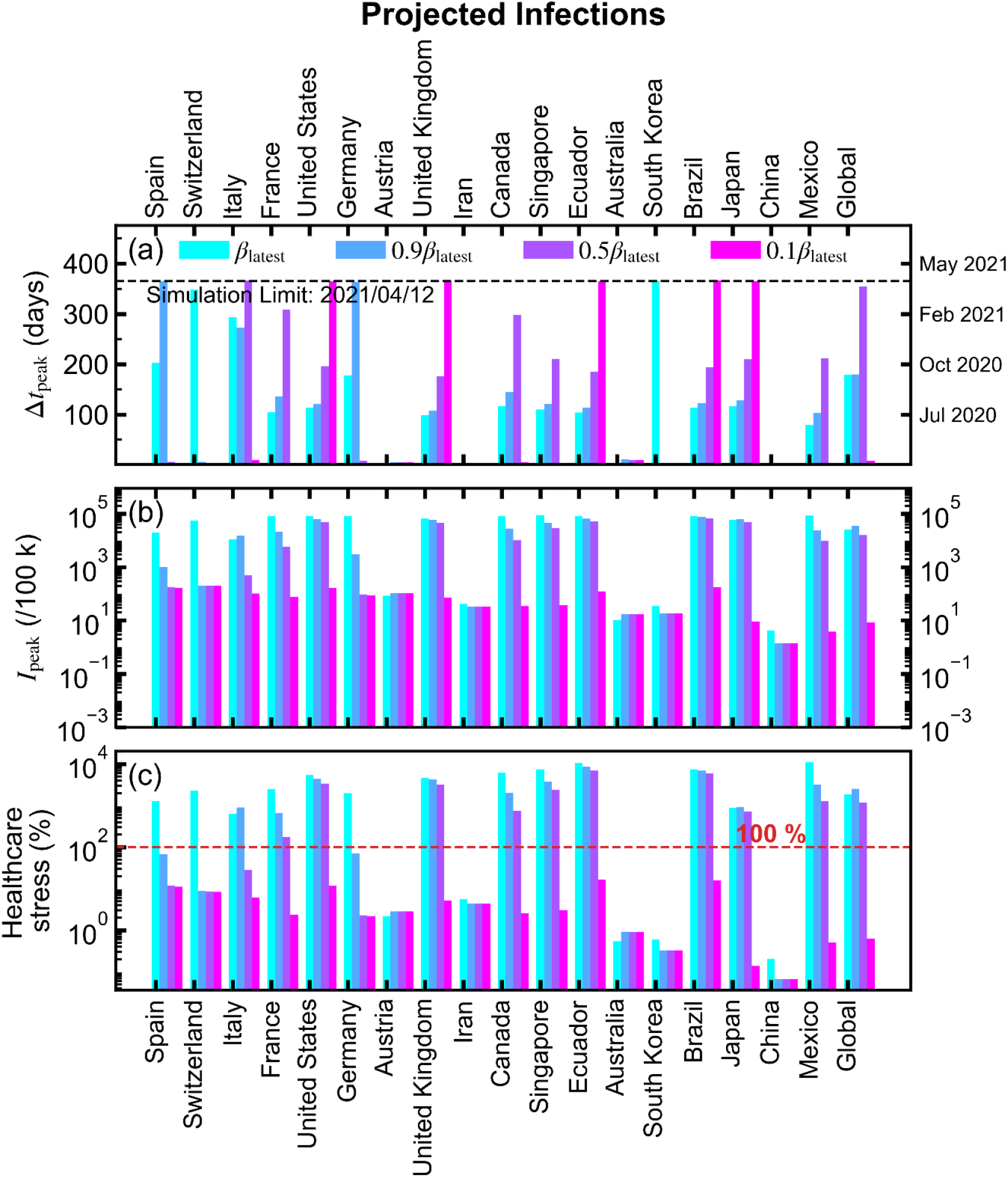
Projected infections for 18 countries using different values of the contagious contact rate (*β*) as related to the effectiveness of the containment. For all figures, cyan color is for the current value of *β* as estimated by our methodology, blue (0.9□), purple (0.5□), and magenta (0.1□). The period estimated until the local peak of the epidemic has been reached is shown in Figure 4a, in which the dashed line indicates the maximum simulation time. Figure 4b shows the total number of cases at the local peak of the epidemic. Figure 4c shows the healthcare stress, defined as the number of cases at the peak requiring hospitalization (20% of the total number of cases) divided by the number of available hospital beds as reported in the World Bank database.

## Discussion

Our multinational modeling of the COVID-19 spread shows a significant heterogeneity of the transmission rates across different 18 countries (10% of those reported in JHCRC). Given that the peak in the number of infections is projected to occur at different times across countries (Table 2), it is not feasible to instrument a single policy recommendation that will effectively curb the spread of the epidemic for all countries. For most cases, it is expected that the course of the pandemic will be extended way past the third quarter of 2020, when the number of new cases is projected to decline. Our projections suggest that local measures imposed in most countries so far have not been enough to contain the spread; at least at the time they were implemented. In some scenarios, the likelihood of significantly reducing the number of cases and mortality rates would require tighter measures to reinforce social distancing.^16^ Countries that acted earlier benefited from the social lockdown and those countries facing a more challenging COVID-19 spread should enforce more stringent measures to avoid overwhelming the capacities of their healthcare systems. Should the course of the pandemic continue as it has so far, our model projects that the global peak of confirmed cases will be reached after the third quarter of 2020. It should be bear in mind that, although the date of the peak on the number of infections is projected to occur around the fourth quarter of 2020, the number of active infections and hence the likelihood of becoming infected remains high until the number of contagious individuals drops to a negligible number. This means that containment measures should be extended well beyond the peak of infections, for at least the same period required to reach the peak. Our approach to fit different stages of the COVID-19 spread is useful to evaluate changes in both, government response and adherence to the public health policy.^32^ Notably, our proposed *Q*_*T*_ metric evidences changes in the contagious contact rates and elucidates country-specific changes in the rate of spread after implementation of measures to contain the virus. So far, social distancing has been implemented differently across countries, perhaps based on local considerations aimed at reducing the socio-economic impact on vulnerable populations.^33^ Preliminary data from China demonstrates that public health interventions including traffic restrictions, social distancing, home quarantine, and universal symptom survey can be temporally associated with a reduced effective reproduction number of SARS-CoV-2.^34^ This fact is in agreement with our data, which shows that China experienced significant reductions in *β* values; indicating a containment of the COVID-19 spread. Austria and South Korea are additional examples of appropriate public health measures, which have effectively curbed the total number of cases.^35,36^ Additional modelling efforts are required to retrospectively assess the effect of social distancing in curbing the pandemic worldwide. The latter likely indicates that countries at early phase of the spread (e.g. Africa) would experience long-term benefits if the governmental policies are adopted before the onset of the exponential progression. Notably, our analysis shows that a larger decrease in the transmission of SARS-CoV-2 is needed to mitigate the healthcare stress (see figure 4c) and decrease the number of total cases. A side result of flattening the peak of infections is that, depending on the local progress of the epidemic, even a stringent reduction in *β* could have the effect of prolonging the time to reach the peak in the number of infections. The relevance of decreasing the contagious contact rate stems from the fact that the lethality rate could be exacerbated once the number of severe and acute cases exceeds the healthcare capacity.^37^ With respect to mobility, the citywide quarantine at Wuhan since January 23, 2020, had negligible effect on the epidemic trajectories for the rest of the country, demonstrating that a 50% reduction in inter-city mobility leads to a negligible effect on epidemic dynamics.^4^ Therefore, we concluded that accounting for inter-city and inter-country mobility in modeling countries where the epidemic is already present is not of paramount importance and could mostly be neglected in the model. Given the heterogeneity of the COVID-19 outbreak, our projections should be helpful in understanding the effect of reducing *β* under different levels of intervention.

We fitted the time series of both confirmed and removed populations, which provides higher confidence in our estimations (see Table 1). However, some limitations to our analysis can significantly influence the model predictions. For instance, our analysis does not consider the fact that the number of confirmed cases could be underestimated (accordingly to official reports). Some countries such as South Korea, Germany and to some extent the US have enacted massive testing which has shown benefits in reporting the spread of the disease,^38^ whilst others countries such as Mexico have enacted a proper sentinel surveillance model because of effectiveness in tracking outbreaks in other respiratory virus infections.^39^ It is noteworthy to mention that using only the official reported numbers our model lessens the effect of methodological biases introduced by curating local reporting approaches. An underestimation in the number of active infections arising from the use of potentially under-sampled data^40^ could lead to offsets in our forecasted peak date and in the expected number of peak infections. A second limitation of our approach is that, in contrast with Bayesian models, no prior distribution for constraining the parameter values to a credible interval is used when fitting the models; this might decrease the precision of the estimations in countries with limited data due to a lag in the pandemic’s spread. Because the model parameters were loosely bounded (see Methods), the basic reproduction number obtained for some countries do not precisely agree with other reports for the early onset (see Supplementary Information). In our study we emphasize the comparison of the contagious contact rate at different stages of the epidemic, based on breaking down the logarithmic progression of the time series using a piecewise fitting methodology, with *R*^*2*^ used as a metric of the goodness-of-fit. This allows us to confidently make comparisons of the progress of the pandemic at different stages and across different countries.

Ongoing data collection and epidemiological analysis are essential parts of assessing the impacts of mitigation strategies against COVID-19. In this study, a multinational SEIR modeling considering three different epidemiological stages was used to compare the COVID-19 spreading dynamics in both global and local scenarios. This allowed us to identify the effectiveness of the containment measures across 18 countries and determine the potential burden to the healthcare system assuming three different levels of reduction of the contagious contact rate. Our estimations for the global progress of COVID-19 indicate that the maximum number of active infections will take place after the third quarter of 2020. However, a few countries will experience a decline within the coming weeks. As the contagious contact rate decreases, we observe a monotonic decrease in the peak number of infections. Nevertheless, it should be noted that, depending on the local state of the spread, a side effect of flattening the peak of infections is that some countries will experience a considerable delay of the evolution of COVID-19. For all studied countries, a 90% reduction in the contagious contact rate is required to avoid overwhelming the healthcare capacity. COVID-19 exhibits a heterogeneous spread, a fact that can be related to several variations in both timing and efficacy of the containment measures implemented in each country. Our results provide informative guidelines to avoid healthcare overcapacity and weigh the long-term effects of more lenient policies.

## Data Availability

The authors confirm that the data supporting the findings of this study are available within the article and/or its supplementary materials.

https://github.com/erickmartinez/covid19

## Contributors

EML and FFCT conceived the study; EML and FFCT designed the model; EML programmed the SEIR model; EML, FFCT, JJN and OYBC analyzed the data; JJN, OYBC, EML and FFCT interpreted the projections; EML, FFCT, JJN and OYBC wrote original paper. All authors made data curation and resources, and approved the final version for submission.

## Source code

Python sources for this work are available at https://github.com/erickmartinez/covid19

## Supplementary results

**S1**. China

**S2**. Europe

**S3**. Asia and Australia

**S4**. Middle East

**S5**. North and South America

**S6**. Basic reproductive number estimations

